# One-Year Follow-up of Young People with ME/CFS Following Infectious Mononucleosis by Epstein-Barr Virus

**DOI:** 10.1101/2023.07.24.23293082

**Authors:** Rafael Pricoco, Paulina Meidel, Tim Hofberger, Hannah Zietemann, Yvonne Mueller, Katharina Wiehler, Kaja Michel, Johannes Paulick, Ariane Leone, Matthias Haegele, Sandra Mayer-Huber, Katrin Gerrer, Kirstin Mittelstrass, Carmen Scheibenbogen, Herbert Renz-Polster, Lorenz Mihatsch, Uta Behrends

## Abstract

**Background:** Infectious mononucleosis, caused by the Epstein-Barr Virus (EBV-IM), has been linked to the development of myalgic encephalomyelitis/chronic fatigue-syndrome (ME/CFS) in children, adolescents, and young adults. Our study presents the first cohort of young individuals in Germany who were diagnosed with ME/CFS following EBV-IM.

**Methods:** We conducted a one-year follow-up of 25 young people diagnosed with ME/CFS at our specialized tertiary outpatient service by clinical criteria requiring post-exertional malaise and with documented EBV-IM as the triggering event. Demographic information, laboratory findings, frequency and severity of symptoms, physical functioning, and health-related quality of life (HRQoL) were assessed at first visit as well as 6 and 12 months later at follow-up visits.

**Results:** The physical functioning and HRQoL of the cohort were significantly impaired, with young adults displaying more severe symptoms, as well as worsening of fatigue, physical and mental functioning, and HRQoL throughout the study, compared to adolescents. After one year, we found that 6/12 (54%) adolescents no longer met the diagnostic criteria for ME/CFS, indicating partial remission, while all young adults continued to fulfill the Canadian consensus criteria. Improvement in children was evident in physical functioning, symptom frequency and severity, and HRQoL, while young adults had little improvement. EBV serology and EBV DNA load did not correlate with distinct clinical features of ME/CFS, and clinical chemistry showed no evidence of inflammation. Remarkably, the median time from symptom onset to ME/CFS diagnosis was 13.8 (IQR: 9.1–34.9) months.

**Conclusions:** ME/CFS following EBV-IM in young people is a severely debilitating disease with diagnoses protracted longer than one year in many patients and only limited responses to conventional symptom-oriented medical care. Although younger children may have a better prognosis, their condition can fluctuate and significantly impact their HRQoL. Our data emphasize that biomarkers and effective therapeutic options are also urgently needed for this very young age group to better manage their medical condition and pave the way to recovery.

## 1 Introduction

Myalgic encephalomyelitis/chronic fatigue syndrome (ME/CFS) is a complex and debilitating multi-system disease characterized by fatigue and post-exertional malaise (PEM) together with additional symptoms, including unrefreshing sleep, cognitive impairment, orthostatic intolerance, and/or chronic pain. It has a profound impact on all aspects of daily life. Up to 25% of patients are severely affected and bound to home or bed [1, 2]. ME/CFS has been identified as an important cause for long-lasting school absence [3–8] and results in a significant reduction of health-related quality of life (HRQoL) [7, 9–11].

Although ME/CFS is not rare, it is still not well recognized by doctors, teachers, and the general population. Pre-pandemic global prevalence estimates for ME/CFS were 0.3–0.5%, with two age peaks at onset of 11–19 and 30–39 years [12]. The prevalence reported for children and adolescents ranged from 0.1% to 1.9%, depending on the case definition, geographical region, and screening method. A study from the US indicated that up to 95% of children with ME/CFS may remain undiagnosed [13]. The post-pubertal female-to-male ratio is 3–4:1 [13, 14], thus adolescent girls are predominant amongst ME/CFS patients in pediatrics.

Infectious diseases are the most frequent triggers of ME/CFS, accounting for 23–90% pediatric cases [4, 15–17]. Of a large pediatric cohort from Australia 80% of patients recalled an initial infection, and 40% a triggering infectious mononucleosis by Epstein-Barr virus (EBV-IM) [7]. ME/CFS was reported in 13%, 7%, and 4% of 301 adolescents in the US at 6, 12, and 24 months after the onset of EBV-IM [18, 19], respectively, and in 23% of 238 college students at 3–6 months after EBV-IM. 8% of these college students were severely affected [20]. While EBV was recognized as the most prominent viral trigger of ME/CFS until 2019 [7, 19–31], it became outranked by severe acute respiratory coronavirus type 2 (SARS-CoV2), which was estimated to cause at least a doubling of ME/CFS cases worldwide and in Germany [32–34].

The pathomechanisms of ME/CFS and PEM are not yet fully understood. Genetic polymorphisms might contribute to pathogenic immune dysregulation after infections [35]. Evidence is increasing that immunological and vascular changes might result in hypoperfusion of muscles and brain [36]. Microbiome dysbiosis, defects in energy metabolism, dysregulated hormones, and vagus nerve dysfunction have been discussed [21, 37–39]. A causative role of herpes virus reactivation was evaluated but has not been proven yet [40–45]. However, we recently reported, that EBV might as well trigger pathogenic autoimmunity by molecular mimicry [46].

Several risk factors have been suggested for ME/CFS after EBV-IM, including disease severity and days-in-bed during the acute phase, initial pain and autonomic symptoms, lower mental health scores, higher scores for anxiety, depression, and perceived stress, female gender, as well as distinct laboratory findings (e.g. elevated CRP and cytokine levels). However, different case definitions have been used and findings were inconsistent [20, 28–30, 47]. Jason and colleagues found that baseline anxiety, stress, depression, or coping skills did not predict the development of ME/CFS after EBV-IM, while preceding symptoms of the ME/CFS spectrum before EBV-IM increased the risk [20].

ME/CFS is diagnosed according to clinical criteria and after exclusion of other diseases that might explain the symptoms [48]. In adults the systemic exertion intolerance disease (SEID)/Institute of Medicine (IOM) criteria [49] are recommended for screening and the Canadian Consensus Criteria (CCC) [50] for diagnosis and research. For children and adolescents the CCC were adapted in a “pediatric case definition” (here abbreviated as PCD-J) by Jason and colleagues [51] and modified in a “clinical diagnostic worksheet” (here abbreviated as CDW-R) developed by Rowe and colleagues who thereby referred to less typical pediatric cases [6]. All four scores require PEM as an essential criterion. The interdisciplinary diagnostic workup addresses not only differential diagnoses but also possible comorbidities such as autoimmune thyroiditis, orthostatic intolerance, or hypermobile Ehlers Danlos (hEDS) syndrome, and includes a psychological evaluation. A 10-minute standing test (NASA lean test) or tilt table test is recommended to diagnose postural orthostatic tachycardia syndrome (PoTS) [6, 48] which itself is often triggered by infections.

As of now, no specific treatment is available for ME/CFS. A consequent self-management with pacing is recommended, and can be supplemented by various relaxation strategies, and sleep hygiene. Symptom-oriented, complex medical support is essential to reduce the severity and frequency of symptoms, and usually includes non-pharmaceutical and pharmaceutical approaches. Psychosocial support may help with implementing pacing and coping strategies, and could achieve adequate measures for education and home care [6, 48, 52]. Experimental strategies targeting the immune, vascular, and nervous system seem promising [53].

With adequate treatment, the course of disease seems to be better in children and adolescents compared to adults, with pediatric recovery rates of 5–83% [6–8, 15–17, 27, 54–61]. Recovery rates in young people have been operationalized by measuring school attendance, symptom frequency and severity, as well as fulfillment of diagnostic criteria at clinical visits or via self-report [55, 62]. In an Australian pediatric cohort, one and two thirds of the patients recovered after 5 and 10 years, respectively, with a median disease duration of 5 (1–15) years in those who recovered [7].

However, in many ME/CFS patients the course of disease is fluctuating, with periods of deterioration (“crashes”), stabilization, improvement, or relapse-remitting cycles [6, 7, 23]. About 40% of adult patients are estimated to improve over time, but only 5% fully recover [63, 64]. Inferior outcomes might in part be due to inappropriate management resulting from inadequate disease- specific knowledge of medical staff [3, 54, 65, 66], a lack of medical services and barriers to the health care system for patients with ME/CFS [67–69], and stigmatization of patients.

Here we present a first cohort of young ME/CFS patients diagnosed after documented EBV-IM at our MRI Chronic Fatigue Center for Young People (MCFC) and recruited into our prospective MUC-CFS studies for pediatric and adult patients. The MCFC, so far, is the only pediatric university center in Germany specialized on the diagnosis and treatment of children and adolescents with ME/CFS. The MUC-CFS studies provide detailed information on patient demographics, disease phenotypes, and quality of life at the time of diagnosis, as well as during follow-up visits at 6 and 12 months. Our primary objective was to assess disease trajectories in these young individuals and to explore potential differences between children and young adults within the cohort.

## 2 Methods

### 2.1 Study Design and Diagnostic Work-up

This was a single-center prospective cohort study conducted at a tertiary pediatric university hospital in Munich, Germany. Adolescents and adults until and including the age of 25 years who were diagnosed with ME/CFS after EBV-IM were reassessed at 6 and 12 months by the interdisciplinary team of the MCFC between March 2019 and November 2022.

Documented EBV-IM was defined as a combination of typical symptoms (e.g., fever, fatigue, sore throat, lymphadenopathy, and/or splenomegaly) and typical serology (positive IgM and/or IgG antibodies against EBV viral capsid antigen (VCA) without IgG antibodies against EBV nuclear antigen 1 (EBNA-1), and – in some cases – with documented subsequent EBNA-1-IgG seroconversion).

ME/CFS was diagnosed by a multidisciplinary board. Diagnostic criteria were used depending on age: For adults (≥ 18 years) the CCC were used, adolescents had to fulfill either the CCC with a disease duration of at least three months or the less strict criteria of the CDW-R with a disease duration of at least six months. In both cases, PEM had to last for more than 14 hours after mild exertion. Since ME/CFS is a diagnosis of exclusion, all patients underwent thorough medical examinations (laboratory analyses, ECG, UCG, EEG, cMRI, and pulmonary function analyses), including a psychological evaluation. Depending on the clinical presentation supplementary investigations were implemented. Laboratory testing was performed as recommended by Rowe and colleagues [6], and a 10-minute standing test (NASA lean test) was conducted to screen for orthostatic intolerance (OI), PoTS, or orthostatic hypotonia (OH).

### 2.2 Standard of Care/Treatment

All patients received a comprehensive, symptom-oriented treatment, the latter of which included both pharmaceutical and non-pharmaceutical interventions to manage symptoms such as pain, sleep disorder, OI and/or psychological comorbidities. They were provided with extensive guidance on self-management and received recommendations for psychosocial support, including adequate measures for adapted school education and home care.

### 2.3 Ethical Consideration

Patients and parents (of patients < 18 years) provided informed consent prior to inclusion. The study was approved by the local Ethics Committee (529/18 S-KK, 485/18 S-KK) and conducted in accordance with the Declaration of Helsinki and its later amendments.

### 2.4 Data Collection

#### 2.3.1 Clinical Data

Data were collected from standard clinical records and additional questionnaires from 2019 to 2022. For follow-up visits, questionnaires were mailed to the families one month before a personal or telephone visit. Telephone visits took place when personal visits were not medically indicated, and ME/CFS criteria (CCC for all and CDW-R for adolescent patients) were evaluated.

#### 2.3.1 Patient Reported Outcome Measures

The <u>Pediatric Quality of Life Inventory (PedsQL</u>) was utilized to assess HRQoL in pediatric patients. It compromises 20 items and four subscales, namely physical, emotional, social, and school functioning, and has been found to have good internal consistency and reliability [70].

The <u>Short Form-36 Health Survey (SF-36)</u> was administered to all patients to measure HRQoL. It is a 36-item questionnaire that consists of eight subscales (physical functioning, role physical, general health, bodily pain, social functioning, vitality, role emotional, and mental health). Lower scores on each subscale indicate more impairment. The SF-36 has been well-validated and has good internal consistency and discriminate validity. It was designed for children older than 13 years [71]. Younger patients were provided with an in-house supplementary for additional explanations if necessary. Score results are presented as mean ± standard deviation (SD).

The <u>Chalder Fatigue Scale (CFQ)</u> was used as a validated instrument to measure physical and mental fatigue. The scale consists of 11 items, with responses rated on a Likert scale from 0–3. The total score reflects the sum of responses, with a score of 33 indicating most severe fatigue [72]. Score results are presented as mean ± SD.

The <u>Charité Symptom Inventory</u> was adapted from the CDC Symptom Inventory and rated frequency and severity of typical symptoms of ME/CFS within the month prior to the visit. Scales rated from 0 (not present) to 3 (severe) for the severity and from 0 (not present) to 4 (always) for the frequency of symptoms [73].

The <u>Bell Score</u> is a short and widely used instrument to assess the severity of ME/CFS [74]. Adolescent patients received a modified version with adapted wording (e.g. “school” instead of “work”).

### 2.4 Statistical Analysis

The statistical analysis was conducted using R Version from 4.2.1 (“Funny-Looking Kid”) [75]. Categorial variables were compared between groups using Fisher’s exact test or Pearson’s chi-squared test, as appropriate. For numeric variables, we used the Wilcoxon rank-sum test or Kruskal-Wallis test, as appropriate. Correlation analysis was performed using Spearman’s rank correlation coefficient. Cox regression analysis was used to assess the association between independent variables and the time-to first presentation in the MCFC. Repeated measures correlation were used to measure within-subject correlation of patient reported outcome measures [76]. Given the small sample size and lack of adjustment for multiple testing, all P-values were considered exploratory. Level of significance was set to α = 0.05.

## 3 Results

### 3.1 Baseline Demographics and Clinical Characteristics of the Cohort

Baseline characteristics are shown in **Table 1**. All 25 patients (80% female) had a history of EBV-IM and documented serological findings, indicating EBV primary infection at the time of disease onset. There were no significant differences between adolescents (48%, median age at onset 15, IQR 13 –15) and young adult (≥ 18 years) patients (52%, median age at onset 10, IQR 18 – 21) concerning demographics, medical and family history, and current medical care. The median time between EBV-IM and ME/CFS diagnosis at our center was 13.8 months (range 4–84) with no significant difference between males and females (P = 0.272), or adult and adolescent patients (P = 0.596). The time delay from symptom onset to diagnosis was less than 6, 12, and 24 months in 1/13 (8%), 5/13 (38%), and 7/13 (54%) adult patients as well as in 1/12 (8%), 5/12 (42%), and 10/12 (83%) adolescent patients (**Supplementary Figure S1**).

**Table 1.** Baseline Demographics and Clinical Characteristics of the Cohort.

| Characteristics | All | Adults | Adolescents | p-value<sup>1</sup> |
| --- | --- | --- | --- | --- |
| Number of patients | n=25 | n=13 | n=12 |  |
| Age and illness duration<sup>2</sup> |  |  |  |  |
| Age at first visit | 18 (16 – 21) | 21 (19 – 22) | 16(14– 16) | <0.001 |
| Age at onset | 16 (14 – 19) | 19 (18 – 21) | 15 (13 – 15) | <0.001 |
| Illness duration in months from onset to first visit | 13.8 (9.1 – 34.9) | 16.9 (9.4 – 44.1) | 13.2 (8.8 – 22.0) | 0.503 |
| Baseline questionnaire results |  |  |  |  |
| Chalder fatigue scale<sup>3</sup> | 25 (5) | 28 (4) | 22 (5) | **0.006** |
| Bell Score<sup>2</sup> | 40 (30 – 50) | 30 (30 – 40) | 50 (40 – 50) | 0.056 |
| SF-36 PCS<sup>3</sup> | 29 (9) | 26 (8) | 32 (9) | 0.151 |
| SF-36 MCS<sup>3</sup> | 42 (11) | 39 (12) | 45 (9) | 0.211 |
| PedsQL<sup>3</sup> | 46 (14) | 35 (11) | 54 (9) | **0.002** |
| Gender<sup>4</sup> |  |  |  |  |
| Female | 20/25 (80) | 11/13 (85) | 9/12 (75) | 0.645 |
| ME/CFS criteria<sup>4</sup> |  |  |  |  |
| CCC | 21/25 (84) | 13/13 (100) | 8/12 (67) | **0.039** |
| CDW-R | 12/12 (100) | N/A | 12/12 (100) | 0.077 |
| Comorbidity<sup>4</sup> |  |  |  |  |
| PoTS | 19/23 (83) | 11/11 (100) | 8/10 (80) | >0.999 |
| Allergies | 12/25 (48) | 7/13 (54) | 5/12 (42) | 0.543 |
| Asthma | 1/25 (4) | 0/13 (0) | 1/12 (8) | >0.999 |
| Neurodermatitis | 1/25 (4) | 1/13 (8) | 0/12 (0) | >0.999 |
| Psychiatric disorder | 3/25 (12) | 2/13 (15) | 1/12 (8) | >0.999 |
| Medical history<sup>4</sup> |  |  |  |  |
| Trauma/surgery | 1/25 (4) | 0/13 (0) | 1/12 (8) | 0.480 |
| Asthma | 2/25 (8) | 1/13 (8) | 1/12 (8) | >0.999 |
| Psychiatric disorder | 1/25 (4) | 0/13 (0) | 1/12 (8) | 0.480 |
| Current medical care<sup>4</sup> |  |  |  |  |
| Complete vaccinations | 22/22 (100) | 11/11 (100) | 11/11 (100) |  |
| Nutrition supplements | 17/24 (71) | 10/12 (83) | 7/12 (58) | 0.319 |
| Prescription medication | 11/24 (46) | 5/12 (42) | 6/12 (50) | >0.999 |
| Degree of disability | 1/25 (4) | 0/13 (0) | 1/12 (8) | >0.999 |
| Medical aid | 0/25 (0) | 0/13 (0) | 0/12 (0) |  |
| Long-term care level | 0/25 (0) | 0/13 (0) | 0/12 (0) |  |
| Family history<sup>4</sup> |  |  |  |  |
| ME/ CFS in family | 2/25 (8) | 1/13 (8) | 1/12 (8) | >0.999 |
| AID in family | 10/25 (40) | 6/13 (46) | 4/12 (33) | 0.688 |
| PID in family | 0/25 (0) | 0/13 (0) | 0/12 (0) |  |
| IM in family | 16/25 (64) | 9/13 (69) | 7/12 (58) | 0.688 |

<sup>1</sup> Fisher's exact test; Wilcoxon rank sum test; Pearson's Chi-squared test
<sup>2</sup> Median (IQR)
<sup>3</sup> Mean (SD)
<sup>4</sup> Number of patients with indicated characteristic/number of patients investigated (%)
CCC, Canadian Consensus Criteria [49], CDW-R, Clinical Diagnostic Worksheet [6]; PoTS, postural orthostatic tachycardia syndrome; AID, autoimmune disease; PID, primary immune deficiency; IM, infectious mononucleosis; PedsQL, Pediatric Quality of Life Inventory; SF-36 PCS, Short Form 36 Health Survey Physical Component Summary Score; SF-36 MCS, Short Form 36 Health Survey Mental Health Component Summary Score. N/A, not applicable.

All adult patients met the CCC and all adolescent patients the CDW-R and/or CCC criteria, as required. Adult and adolescent patients did neither significantly differ concerning the baseline Bell Score nor the SF-36 physical (PCS) and mental health component summary score (MCS). However, adult patients showed significantly higher CFQ scores (adults: 28±4; adolescents: 22±5; P = 0.006), and significantly lower values (adults: 35±11; adolescents: 54±9; P = 0.002) in the PedsQL compared to adolescent patients. All adolescents reported regular absences from school, 2/11 (18%) received some form of complementary home schooling and none reported access to distance schooling. Only one patient had a documented degree of disability, and none received medical care at home.

24/25 (96%) showed comorbidities led by PoTS in 21/23 (83%) and allergies in 12/23 (48%) of all patients. Two patients examined for PoTS did not complete the NASA-lean test due to severe orthostatic symptoms. One patient presented with an anxiety disorder, and two with mixed anxiety and depressive disorder. 17/24 (71%) of the patients took various supplements, and 11/24 (46%) took prescription-only medications, including three patients on antidepressants. 16/25 (64%) patients remembered another family member with a history of EBV-IM, 10/25 (40%) reported a history of autoimmune disease in their family, and 18/25 (72%) either. 2/25 (8%) reported a family member with ME/CFS.

Our patients had consulted several (median 6, range 1–11) private practice doctors across five different specialties (range 1–11) for ME/CFS symptoms. Additionally, 11/20 (55%) had visited at least one hospital. 7/20 (35%) had previously consulted at least one psychotherapist/psychologist. In terms of complementary therapies, 9/20 (45%) patients had sought a naturopath, 6/20 (30%) traditional Chinese medicine, 4/20 (20%) homeopathy, and 5/20 (25%) osteopathy.

### 3.2 Laboratory Findings at Baseline

Laboratory findings at the time of diagnosis were primarily unremarkable, and none significantly differed between adolescent and adult patients (**Table 2**). Besides low vitamin D levels in 14/24 (58%) (range 7–29 ng/ml), the most frequent laboratory findings were elevated antinuclear antibodies (ANA) in 14/25 (56%) (range 1:100–1:800), elevated IgE in 7/25 (28%) and mild anemia in 4/25 (16%) of all patients. 2/6 (33%) adolescents had mildly positive ANA titers (< 1:160), 2/6 (33%) adolescents and 8/8 (100%) adults showed moderately positive titers (1:160–1:640), and 2/6 (33%) adolescents presented with strongly positive titers (≥ 1:640). ANA titers significantly differed between age groups with higher titers in adolescents (P = 0.015). They did not significantly correlate with disease severity (Bell Score: P = 0.452; SF-36 PF: P = 0.858), and were not significantly different between males and females (P = 0.521). High titers were not associated with clinical signs or any other markers of connective tissue disorders. Herpes simplex virus coinfection was not more frequent in adults than in adolescents (P > 0.999). None of the laboratory findings indicated an relevant differential diagnosis. Neither immunoglobulin serum levels nor immunophenotyping of peripheral blood lymphocytes did show any evidence of primary immunodeficiency **(Supplementary Table S1).**

**Table 2.** Selected Laboratory Results at Baseline Visit.

| Laboratory parameter | All<br>n/n (%)<sup>1</sup> | Adults<br>n/n (%)<sup>1</sup> | Adolescents<br>n/n (%)<sup>1</sup> | p-value<sup>2</sup> |
| --- | --- | --- | --- | --- |
| **Blood count** |  |  |  |  |
| Neutropenia (<1,500/ul) | 2/25 (8) | 1/13 (8) | 1/12 (8) | >0.999 |
| Lymphocytes ↑ | 5/25 (20) | 2/13 (15) | 3/12 (25) | 0.645 |
| Thrombocytes ↓ | 0/25 (0) | 0/13 (0) | 0/12 (0) |  |
| Hemoglobin ↓ | 4/25 (16) | 1/13 (8) | 3/12 (25) | 0.322 |
| **Inflammation** |  |  |  |  |
| Sedimentation rate ↑ | 1/21 (5) | 0/13 (0) | 1/8 (12.5) | 0.350 |
| C-reactive protein ↑ | 0/25 (0) | 0/13 (0) | 0/12 (0) |  |
| Ferritin ↑ | 2/23 (9) | 0/13 (0) | 2/10 (20) | 0.178 |
| **Liver function** |  |  |  |  |
| GOT ↑ | 1/25 (4) | 0/13 (0) | 1/12 (8) | 0.480 |
| GPT ↑ | 2/25 (8) | 0/13 (0) | 2/12 (17) | 0.220 |
| Bilirubin ↑ | 1/24 (4) | 1/13 (8) | 0/11 (0) | >0.999 |
| **Immunoglobulins (Ig)** |  |  |  |  |
| IgA ↓↑ | 0/25 (0) | 0/13 (0) | 0/12 (0) |  |
| IgM ↓↑ | 0/25 (0) | 0/13 (0) | 0/12 (0) |  |
| IgG ↓↑ | 0/25 (0) | 0/13 (0) | 0/12 (0) |  |
| IgE ↑ | 7/25 (28) | 3/13 (23) | 4/12 (33) | 0.673 |
| **Infection Serology** |  |  |  |  |
| Cytomegalovirus IgG | 3/25 (12) | 2/13 (15) | 1/12 (8) | >0.999 |
| Herpes simplex virus IgG | 3/25 (12) | 2/13 (15) | 1/12 (8) | >0.999 |
| Toxoplasma IgG | 1/25 (4) | 1/13 (78) | 0/12 (0) | >0.999 |
| Borrelia IgG | 1/25 (4) | 0/13 (0) | 1/12 (8) | 0.480 |
| **Autoantibodies** |  |  |  |  |
| ANA ↑ | 14/25 (56) | 8/13 (62) | 6/12 (50) | 0.561 |
| ANCA ↑ | 1/25 (4) | 1/13 (8) | 0/12 (0) | >0.999 |
| Anti-dsDNA ↑ | 0/25 (0) | 0/13 (0) | 0/12 (0) |  |
| **Endocrinology** |  |  |  |  |
| Cortisol ↓ | 1/25 (4) | 1/13 (18) | 0/12 (0) | 0.480 |
| ACTH ↑ | 1/23 (4) | 0/12 (0) | 1/11 (9) | 0.478 |
| 25-OH-Vitamin-D ↓ | 14/24 (58) | 6/12 (50) | 8/12 (67) | 0.680 |

<sup>1</sup> Number of patients with indicated laboratory parameter/ number of patients investigated (%)
<sup>2</sup> Fisher's exact test; Pearson's Chi-squared test
↑ above normal range; ↓ below normal range; ANA, antinuclear antibodies; ANCA, anti-cytoplasmatic antibodies; anti-dsDNA, anti-double strand DNA

Results from EBV serology, EBV PCR, and EBV IgG immunoblot at baseline visit are displayed in **Table 3**. Adolescents and adults did not show significant differences. Quantitative analyses of EBV viral load using real-time PCR did not detect EBV DNA in the plasma of any patient. 8/20 (40%) patients showed EBV DNA in peripheral blood cells (5/8 very low titers, 1/8 17.7 Geq/10^5, 1/8 70.1 Geq/10^5, and 1/8 121.8 Geq/10^5), and 14/25 (66%) had EBV DNA in throat washes. The detection of EBV DNA in throat washes did not significantly correlate with disease severity (Bell Score: P = 0.686; SF-36 PCS: P = 0.871). All patients showed anti-EBV-VCA IgG as expected, 23/25 (92%) demonstrated anti-EBNA-1 IgG, and 6/25 (24%) anti-EBV-VCA IgM. The detection of anti-EBV-VCA IgM did not significantly correlate with disease severity (Bell Score: P = 0.877; SF- 36 PCS: P = 0.788). Results of EBV immunoblots revealed IgG antibodies against early antigens (EA) p54 and p138, the immediate early antigen BZLF1, virus capsid antigens (VCA) p23 and p18, and the latency antigen EBNA-1 in 8/25 (32%), 5/25 (20%), 18/25 (72%), 23/25 (92%), 24/25 (96%), and 22/25 (88%) patients, respectively.

**Table 3:** EBV Serology, PCR and IgG Immunoblot at Baseline Visit.

| EBV Diagnostics | All<br>n/n (%)<sup>1</sup> | Adults<br>n/n (%)<sup>1</sup> | Adolescents<br>n/n (%)<sup>1</sup> | p-value<sup>2</sup> |
| --- | --- | --- | --- | --- |
| **EBV PCR** |  |  |  |  |
| DNA in cell fraction |  |  |  | 0.927 |
| – | 12/20 (60) | 7/12 (58) | 5/8 (62) |  |
| (+) | 5/20 (25) | 3/12 (25) | 2/8 (25) |  |
| + | 3/20 (15) | 2/12 (17) | 1/8 (12) |  |
| DNA in plasma |  |  |  |  |
| – | 25/25 (100) | 13/13 (100) | 12/12 (100) |  |
| + | 0/25 (0) | 0/12 (0) | 0/12 (0) |  |
| DNA in throat wash |  |  |  | >0.999 |
| – | 11/25 (44) | 6/13 (46) | 5/12 (42) |  |
| + | 14/25 (66) | 7/13 (54) | 7/12 (54) |  |
| **EBV ELISA** |  |  |  |  |
| VCA IgM |  |  |  | 0.110 |
| – | 18/25 (72) | 7/13 (54) | 11/12 (92) |  |
| (+) | 1/25 (4) | 1/13 (8) | 0/12 (0) |  |
| + | 6/25 (24) | 5/13 (38) | 1/12 (8) |  |
| VCA IgG |  |  |  |  |
| – | 0/25 (0) | 0/13 (0) | 0/12 (0) |  |
| + | 25/25 (100) | 13/13 (100) | 12/12 (100) |  |
| EBNA1 IgG |  |  |  | 0.220 |
| – | 2/25 (8) | 0/13 (0) | 2/12 (17) |  |
| + | 23/25 (92) | 13/13 (100) | 10/12 (83) |  |
| **EBV IgG Immunoblot** |  |  |  |  |
| EAp54 |  |  |  | 0.282 |
| – | 14/25 (56) | 7/13 (54) | 7/12 (58) |  |
| (+) | 3/25 (12) | 3/13 (23) | 0/12 (0) |  |
| + | 8/25 (32) | 3/13 (23) | 5/12 (42) |  |
| EAp138 |  |  |  | 0.233 |
| – | 14/25 (56) | 6/13 (46) | 8/12 (67) |  |
| (+) | 6/25 (24) | 5/13 (38) | 1/12 (8) |  |
| + | 5/25 (20) | 2/13 (15) | 3/12 (25) |  |
| BZLF1 |  |  |  | 0.293 |
| – | 3/25 (12) | 3/13 (23) | 0/12 (0) |  |
| (+) | 4/25 (16) | 2/13 (15) | 2/12 (17) |  |
| + | 18/25 (72) | 8/13 (62) | 10/12 (83) |  |
| VCAp23 |  |  |  | >0.999 |
| – | 2/25 (8) | 1/13 (8) | 1/12 (8) |  |
| (+) | 0/25 (0) | 0/13 (0) | 0/12 (0) |  |
| + | 23/25 (92) | 12/13 (92) | 11/12 (92) |  |
| VCAp18 |  |  |  | 0.480 |
| – | 1/25 (4) | 0/13 (0) | 1/12 (8) |  |
| (+) | 0/25 (0) | 0/13 (0) | 0/12 (0) |  |
| + | 24/25 (96) | 13/13 (100) | 11/12 (92) |  |
| EBNA-1 |  |  |  | 0.344 |
| – | 2/25 (8) | 0/13 (0) | 2/12 (17) |  |
| (+) | 1/25 (4) | 1/13 (8) | 0/12 (0) |  |
| + | 22/25 (88) | 12/13 (92) | 10/12 (83) |  |

<sup>1</sup> Number of patients with indicated laboratory parameter/ number of patients investigated (%)
<sup>2</sup> Fisher's exact test
EBV, Epstein-Barr Virus; VCA, virus capsid antigen; EBNA, EBV nuclear antigen; EA, early antigen.

### 3.3 ME/CFS Criteria at Follow-up Visits

Follow-up data were available at 6 months after the diagnosis of ME/CFS from 22/25 (88%) patients, including 10/13 (77 %) adults and 12/12 (100%) adolescents, and at 12 months from 20/25 (80%) patients, including 9/13 (69%) adults and 11/12 (92%) adolescents. Reasons for drop out were recovery (one adolescent), worsening of symptoms (one adult) and unknown (two patients).

The changes in CCC and CDW-R criteria fulfilment are visualized in **Fig. 1**. Of the adult patients, seven fulfilled the CCC at all three visits. One became CCC negative at 6 months but met the CCC criteria again at 12 months (**Fig. 1A**). Six adolescent patients were still positive for the CDW-R criteria at 6 months and only four at 12 months follow-up. One patient became CDW-R negative at 6 months but met the CDW-R criteria again at 12 months (**Fig. 1B**). By 6 months one and by 12 months three additional pediatric patients had turned 18 years old, and therefore the CDW-R criteria were not applicable anymore (indicated by N/A in **Fig. 1C**). The CCC criteria were fulfilled by 8/12 (67%) of the adolescents at the first visit, by 4/12 (33%) at the 6-month visit and by 4/11 (36%) at the 12-month follow-up. Two adolescent patients who were CCC positive at 12 months had been negative at the previous visits (**Fig. 1C**). At the 6-month follow-up, 7/12 (58%) adolescents met either the CCC or the CDW-R criteria, and 5/11 (45%) met either of both at the 12-month follow-up (**Fig. 1D**). The five patients who did not fulfill CCC or CDW-R criteria anymore, still fulfilled some domains of the CCC or CDW-R criteria and therefore showed only partial remission of postviral disease. For two patients the only persisting symptom was OI. One patient reported fatigue with limitations in daily life and headaches. Three patients had several persisting symptoms but did not complain of fatigue with limitations in daily life anymore.

**Figure 1.**
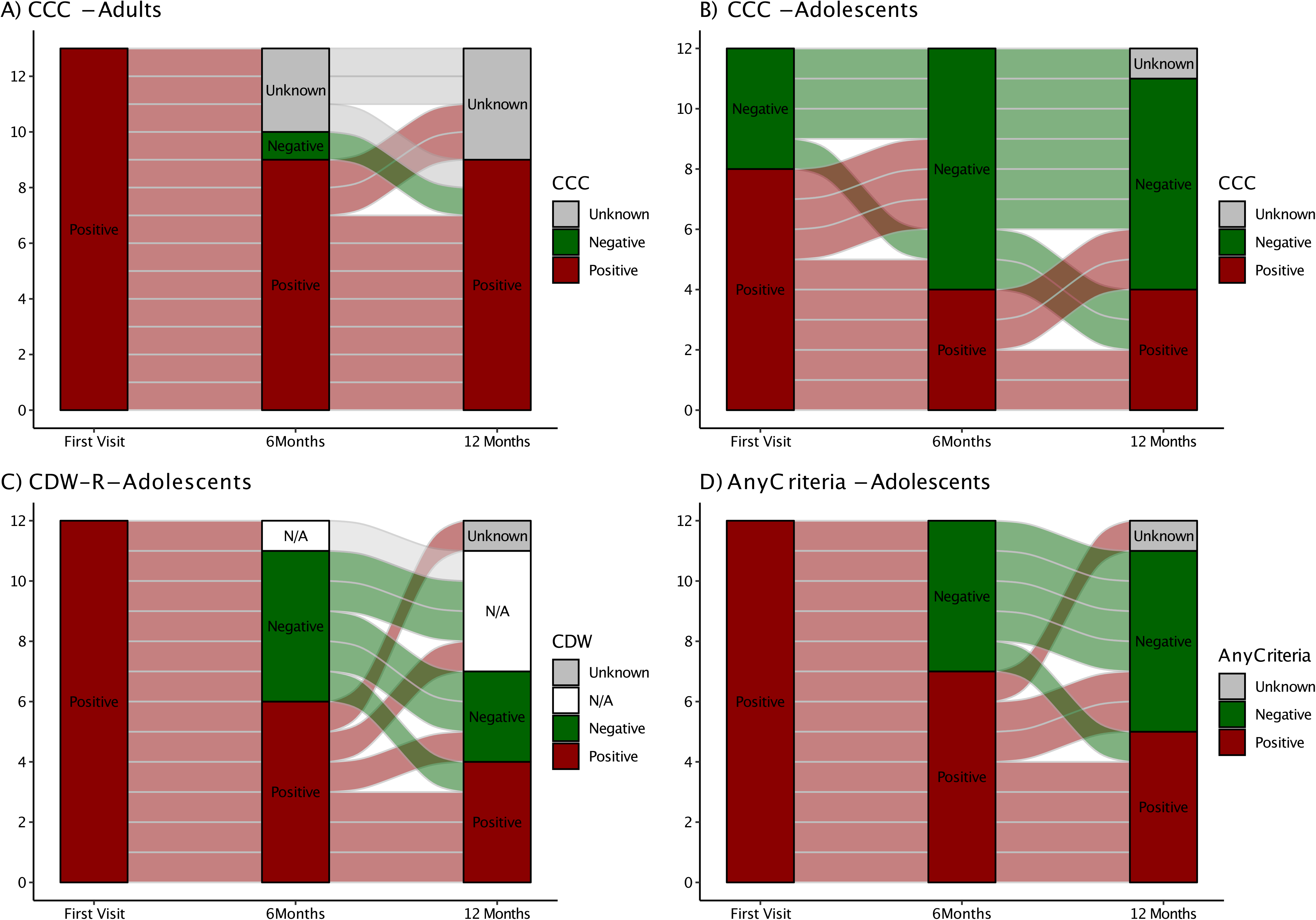
Alluvial chart illustrating ME/CFS diagnostic criteria fulfillment over the study period for adults and adolescents. The chart depicts diagnostic criteria fulfillment (red) or non-fullfillment (green) at the first visit and at visits 6 and 12 months later. (Non-)fullfillment of Canadian Consensus Criteria (CCC) is shown for adults. (Non)-fullfillment of CCC only, Rowe’s diagnostic worksheet (CDW-R) criteria only, or either of both (CCC or CDW-R) is shown for adolescents. CDW-R criteria were not applicable anymore (N/A) when adolescent patients had turned 18 years. Overall, the chart provides an overview of the changes in diagnostic status for ME/CFS patients over time.

All patients in partial remission were adolescents (P = 0.005) and had a relatively short illness duration of less than three years (mean 24 months, range 15–34 months). They had significantly less fatigue (CFQ Likert score: P = 0.001) and higher HRQoL (PedsQL: P = 0.026) at diagnosis, compared to patients without partial remission (**Supplementary Table S2**). Further, patients in partial remission did not significantly differ in any of the other baseline characteristics and laboratory parameters tested, including EBV serology, EBV PCR or EBV IgG Immunoblot (**Supplementary Table S2 and S3**).

### 3.4 Number, Frequency, and Severity of Symptoms at Follow-up Visits

At the baseline visit, patients presented with 27± 5 symptoms in mean (±SD), with 15±5 occurring at least frequently (**Fig. 2**). The most common symptoms reported at least frequently (3 or 4 on Likert scale) were fatigue (96%), performance limitations in daily life (96%), need for rest (92%) and PEM (83%). The most common severe (3 on Likert scale) symptoms were post-exertional malaise (46%), stress intolerance (38%), fatigue (33%), performance limitations in everyday life (33%) and unrefreshing sleep (33%). The number of symptoms and the severity and frequency of individual symptoms did not significantly change between the baseline and two follow-up visits (**Supplementary Table S4**). Adults reported slightly more symptoms (29±3) than adolescents (25±7, P = 0.084). Symptoms occurring at least frequently were more common in adults than adolescents (19±6 vs. 12±3, P = 0.006). This difference was also evident at the two follow-up visits **(Supplementary Table S5)**.

**Figure 2.**
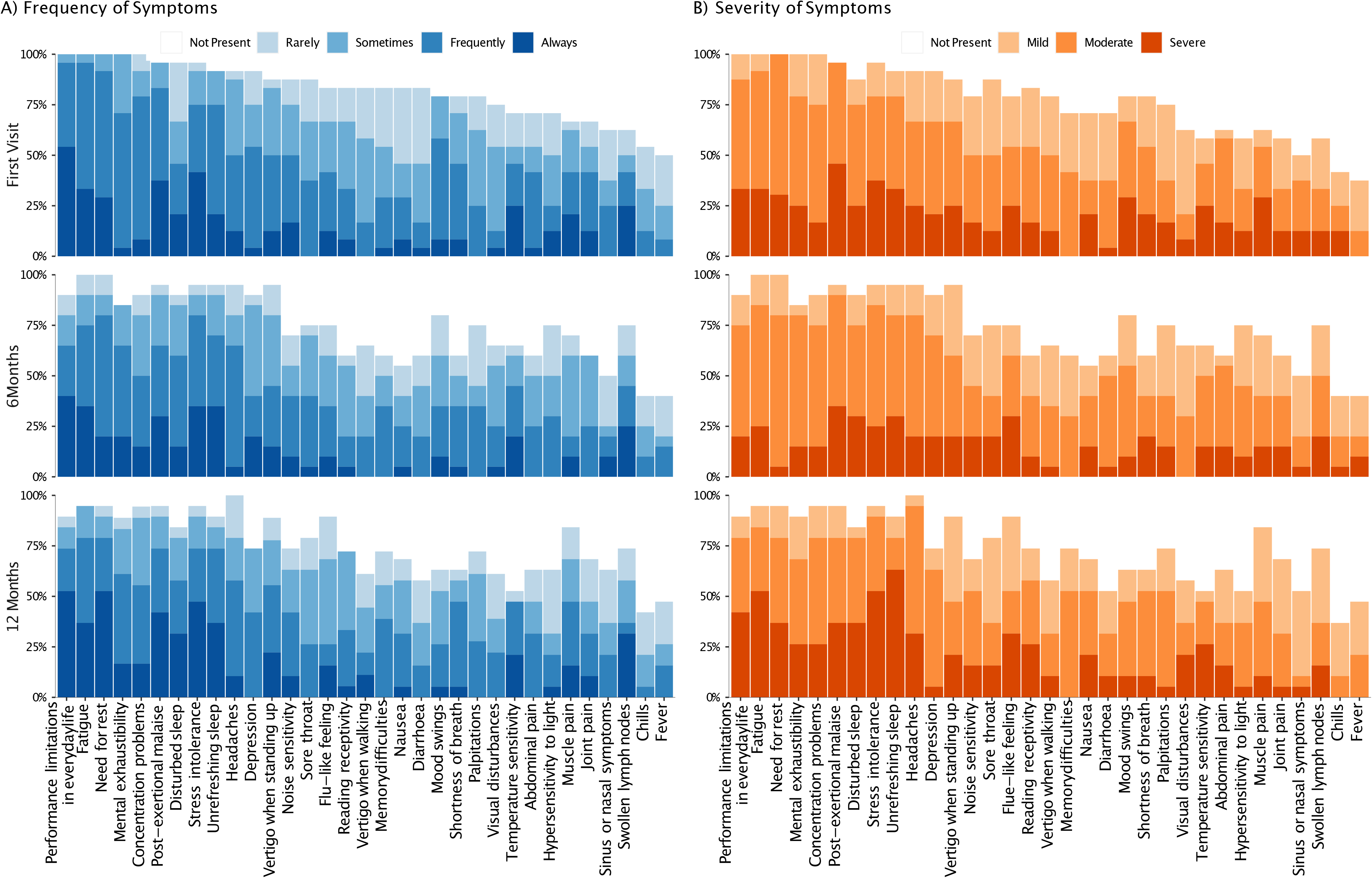
Frequency and severity of symptoms at diagnosis (first visit) and at visits six and twelf months later The bar-chart displays individual symptoms on the x-axis, while the y-axis shows the frequency of symptoms on the left and the severity of symptoms on the right. The severity scale for each individual symptom ranged from 0 (not present) to 4 (severe), and the frequency scale from 0 (not present) to 5 (always present). At each time point, the chart shows the proportion of patients reporting each symptom with respective severity and frequency rating, indicated by color-code. The chart provides an overview of the most common symptoms in ME/CFS patients, how frequently they occured, and how severe they were. It also illustrates how the frequency and severity of symptoms changed over time, with some symptoms improving or worsening and others remaining consistent.

### 3.5 Patient-Reported Outcome Measures

#### 3.4.1 Chalder Fatigue Scale

Fatigue was further characterized by the CFQ. At the first visit, the patients’ CFQ Likert score was 25±5, and did not significantly change at the follow-up visits. While adult patients showed a moderate worsening from the first (28±4) to the follow-up visits (6-months: 28±4; 12-months: 29±4), adolescent patients showed a moderate improvement from the first (22±5) to the follow-up visits (6- months: 19±9; 12-months: 18±9) (**Table 4**, **Fig. 3A**). At all visits, adolescent patients had significantly less fatigue than adults (first visit: P = 0.006; 6-months: P = 0.016; 12 months: P = 0.003) (**Supplementary Table S6**).

**Figure 3.**
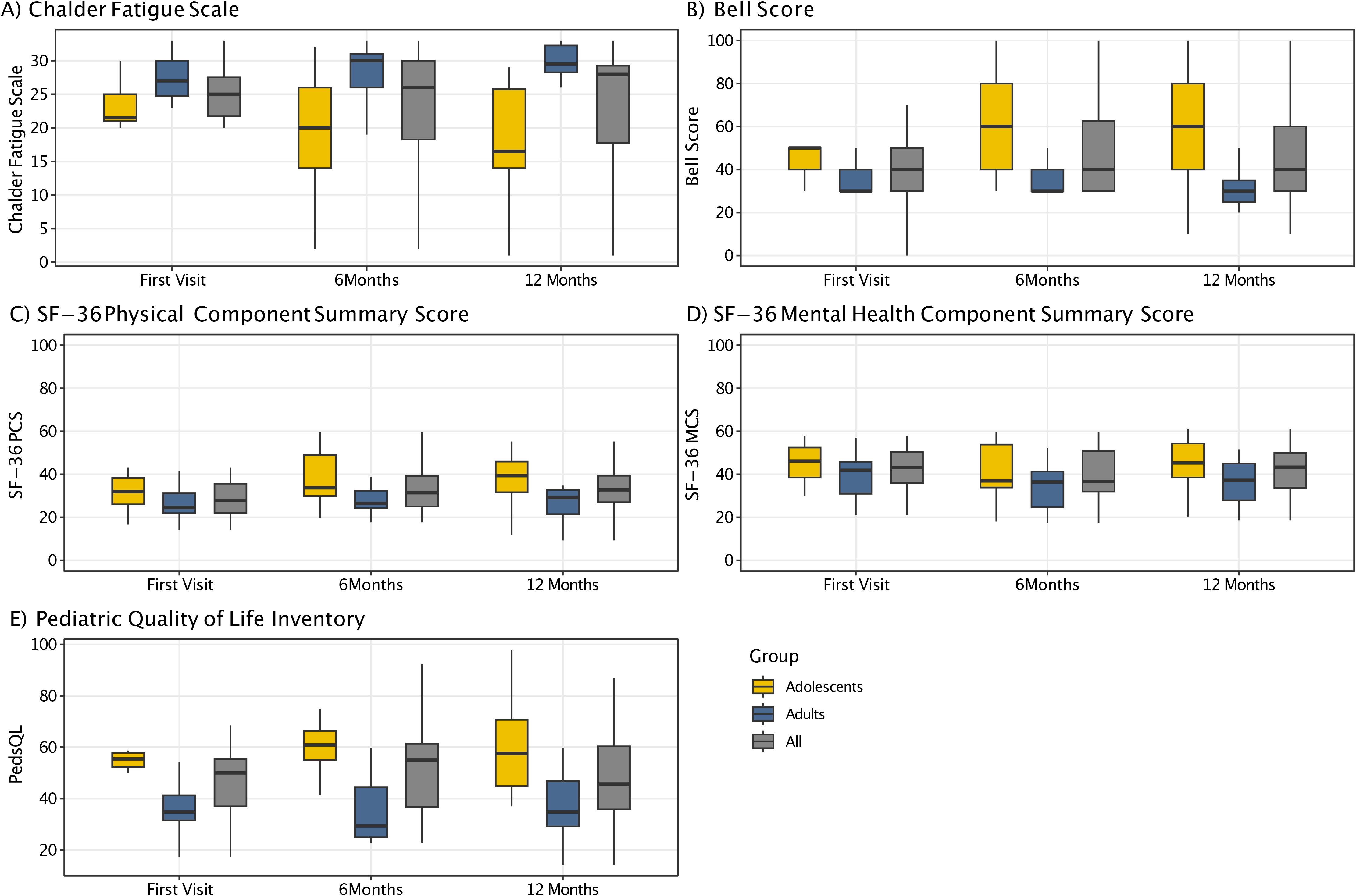
Patient reported outcome measures at diagnosis (first visit) and at visits six and twelf months later Boxplots displaying the dynamics of results from the Chalder Fatigue Scale (A), the Bell Score (B), the SF-36 physical (C) and mental health component summary score (D), and the Pediatric Quality of Life Inventory (E) for the entire cohort as well as for adolescents and adults only, respectively.

**Table 4.**
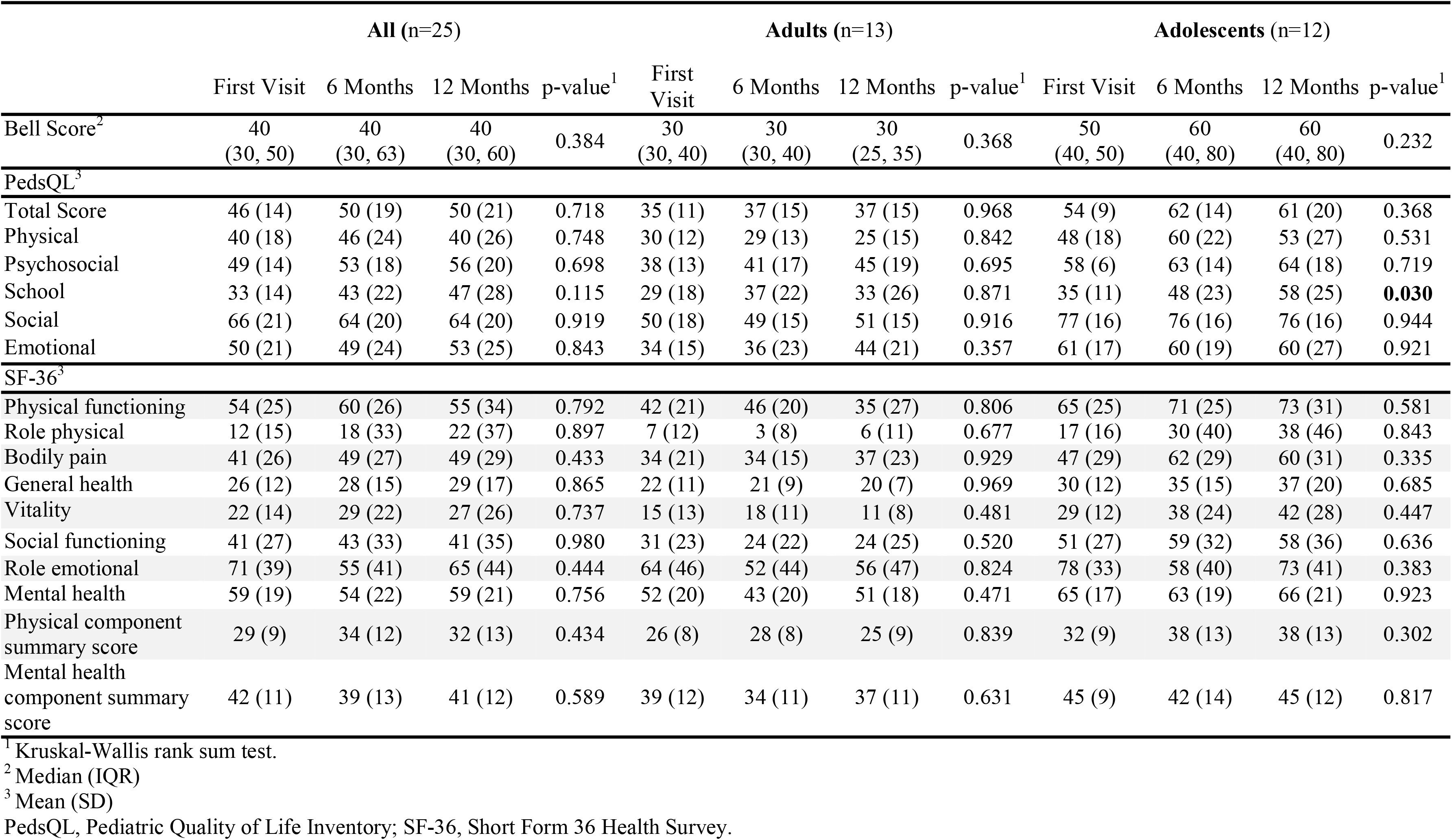
Patient-reported Outcome Measures of the Cohort.

#### 3.4.2 Bell Score

The median Bell Score was 40 (IQR: 30–50) and did not significantly change at the follow-up visits (P = 0.384). Adults had a median Bell Score of 30 at all visits. Adolescents’ Bell Score moderately, but not significantly, improved from the first (median: 50, IQR: 40–50) to the 6-month and 12-month follow-up visits (both median: 60, IQR: 40–80) (P = 0.232) (**Table 4**, **Fig. 3B)**. The Bell Score of adolescents was significantly better than adults’ Bell Score at all visits (first visit: P = 0.019; 6 months: P = 0.019; 12 months: P = 0.007) (**Supplementary Table S6**).

#### 3.4.3 Short Form-36

SF-36 summary and subscales did not significantly change from the first to the two follow-up visits, neither for all nor for adult or adolescent patients separately. However, adolescents had a significantly better physical component summary score at the 12-month visit than adults (P = 0.013) (**Table 4**, **Fig. 3C and 3D**). At the first visit adolescents were significantly better concerning physical functioning (P = 0.039) and vitality (P = 0.012), at the 6 months visit at physical functioning (P = 0.039), pain (P = 0.039), general health (P = 0.032), social (P = 0.025), and mental health (P = 0.025), and at the 12 months visit at physical functioning (P = 0.019) and vitality (P = 0.010). There was no significant difference in the mental health component summary score at none of the visits (**Supplementary Table S6**). In the self-perceived health transition 6/10 (60%) adolescents and none of the adults rated their general health at least somewhat better than in the previous year at the 12 months visit (**Supplementary Table S7**).

#### 3.4.4 Pediatric Quality of Life Inventory

The PedsQL total score did not significantly change from the first to the two follow-up visits. The subscale scores were lowest for the school functioning and physical functioning domains, and highest for social functioning. School functioning in adolescents was the only subscale that significantly increased over time (P = 0.03) (**Table 4**, **Fig. 3E)**. Except for the school subscale at the first 6-month visit and emotional subscale at the 12-months visit, all subscales were significantly different at all visits between adults and adolescents (**Supplementary Table S6**).

For all patients the greatest correlation between different patient-reported outcome measures (CFQ, Bell Score, PedsQL, SF-36) was between CFQ and PedsQL (r = -0.76, P < 0.001), indicating that more severe fatigue was associated with lower HRQoL (**Figure 4**). The most prominent difference between adults and adolescents was that CFQ and Bell Score correlated significantly for adolescents but not for adults (adults: r = -0.29, P = 0.209; adolescents: r = -0.77, P < 0.001).

**Figure 4.**
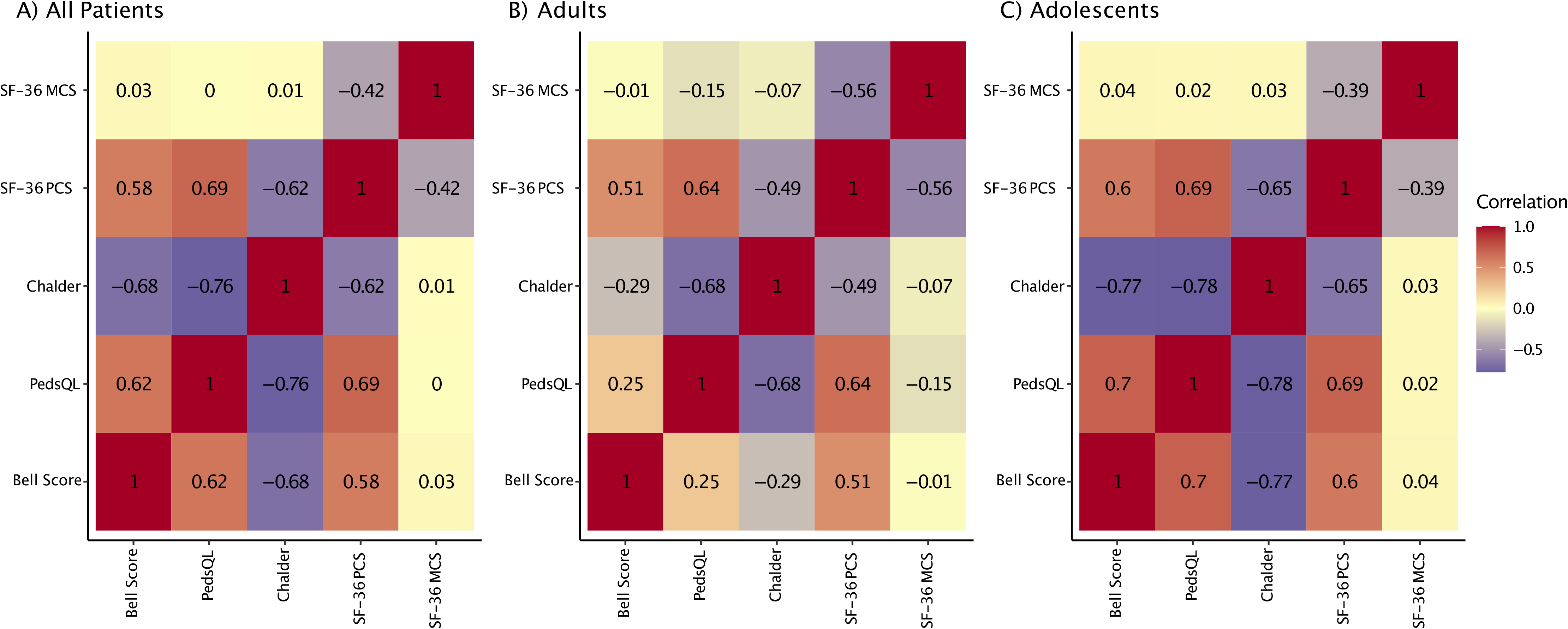
Correlation of patient reported outcomes Heatmap of repeated measures correlations between patient reported outcomes for all patients (A), adults only (B), and adolescents only (C). Repeated measures correlations are a statistical tool to determine the overall within-patient correlation between a pair of variables.

### 3.6 Description of Patients in Partial Remission

At 12 months, results from PROMs for patients in partial remission compared to patients not in partial remission at this time point were median Bell Score 80 (range 40 – 100) versus 40 (range 20 – 80), mean CFQ Score 12.4 (SD 6.7) versus 24.4 (SD 6.2), PedsQL total score 76.1 (SD 16.8) versus 46.6 (SD 15.8), the SF-36 physical health summary score 44.7 (SD 7.7) versus 28.9 (SD 11.5), and the SF-36 mental health summary score 50.9 (SD 7.1) versus 42.9 (SD 10.4). These results indicate that not any more fulfilling the diagnostic ME/CFS criteria does not exclude an ongoing impairment of daily function.

## 4 Discussion

This is the first report on adolescents and young adults with ME/CFS following EBV-IM in Germany. The study provides baseline demographics as well as various clinical data at the initial visit and at two follow-up visits at 6 and 12 months after ME/CFS diagnosis, including diagnostic scores, frequency and severity of symptoms, HRQoL, and physical as well as mental functioning.

ME/CFS is recognized as a debilitating disease which fundamentally impacts social participation, education and HRQoL [6, 7]. However, so far data on ME/CFS in pediatric cohorts have mostly been collected in the US, the UK and Australia, and to our knowledge no pediatric study from Germany is available [6–8, 15–17, 27, 54–61]. While some prospective pediatric studies assessed the occurrence of ME/CFS following EBV-IM [18–20], to our knowledge none evaluated several diagnostic scores requiring PEM longitudinally and/or compared data from adolescents and young adults.

### Baseline Demographics and ME/CFS Diagnosis

Our youngest patient was 14 years-old which was in line with the published ME/CFS age peak at 15–40 years [12, 48, 49]. The observed female predominance (80%) in our cohort is a widely recognized phenomenon in post-pubertal ME/CFS patients with 3–4 times higher prevalence reported in adolescent girls than boys [6, 21, 49].

At the initial visit, all adult patients met the CCC and all pediatric patients met the CDW-R and/or CCC criteria as was required for ME/CFS diagnosis in this study. However, due to the absence of pain (n=3) or neurocognitive manifestations (n=1), 4/12 (33%) of the adolescents did not fulfill the CCC, supporting the use of more sensitive diagnostic criteria for pediatric patients [77]. Different adaptations have been made for diagnosing ME/CFS in children and adolescents [6, 51, 54] of which the CDW-R and Jason’s pediatric case definition (here abbreviated as PCD-J) require PEM as an essential symptom, in line with current recommendations [54, 78]. The largest pediatric ME/CFS follow-up study used the polythetic Fukuda criteria with the modification that the presence of PEM was mandatory [7]. However, most other pediatric follow-up studies used the original polythetic Fukuda criteria or the even broader Oxford criteria for diagnosing ME/CFS [7, 8, 15–17, 27, 55–61] and might thus have included individuals without PEM. Data on the prevalence and clinical outcomes of post-infectious ME/CFS in children and adolescents have thus to be reevaluated. In our ongoing prospective Munich infectious mononucleosis (IMMUC) study [79] as well as the Multicenter Long COVID registry (MLC-R) [80] and the German ME/CFS registry (ME/CFS-R) [81] we are investigating the CCC together with the CDW-R, the PCD-J and the systemic exertion intolerance disease (SEID) criteria, the latter of which have been recommended for diagnosing ME/CFS in any group by the former Institute of Medicine (IOM) [49], the European Network on ME/CFS research (EUROMENE) [48], and the Centers of Disease Control and Prevention (CDC) [82].

In the present study the median diagnostic delay between the onset of EBV-IM and the diagnosis of ME/CFS was more than one year (13.8 months). This is in line with most reports from other countries, indicating long and difficult patient journeys before initial expert consultation or ME/CFS diagnosis of pediatric [3, 65] and adult patients [54, 66]. We did not find any association of gender, age, or disease severity with time to diagnosis according to previous studies [65, 83]. Published reasons for the long diagnostic delay in ME/CFS include little knowledge of families and primary care providers, the requirement for comprehensive differential diagnosis, as well as negative attitudes and beliefs by primary care physicians and psychologists [54, 83, 84]. In Germany, the lack of ME/CFS specialists is most likely contributing to this issue and, due to a higher prevalence of ME/CFS, might affect the adult age-group even more than adolescents and children. Young adults in our study showed a longer prior disease duration compared to children, which might also reflect challenges during transition from pediatric to adult health care services [85].

### Postural Tachycardia Syndrome and Other Comorbidities

In our cohort, only few patients demonstrated any other medical diagnosis prior to the onset of ME/CFS but many showed current comorbidities. The latter included PoTS (83%), allergies (48%), and psychiatric diagnoses (12%). The low prevalence of the latter was in line with other reports on pediatric ME/CFS [6, 86, 87]. In general, comorbidities were more prevalent in studies with adult ME/CFS patients (79–80%) [23, 88].

PoTS has been previously reported in pediatric and adult ME/CFS patients with varying prevalence [6, 54]. 25/26 adolescents with ME/CFS experienced severe orthostatic symptoms associated with syncope (7/25), orthostatic tachycardia with hypotension (15/25), and orthostatic tachycardia without significant hypotension (3/25) during a head-up tilt table test [89]. In adult ME/CFS patients, the prevalence of PoTS was generally reported to be high with reported rates ranging from 5.7% to 70% [49]. The large span of percentages might be due to different cohorts as well as to different PoTS tests and PoTS case definitions [90]. However, PoTS is a frequent post-infectious phenomenon in adolescents and thus, not unexpectedly, a frequent comorbidity of ME/CFS [91]. A timely PoTS diagnosis is important since it impacts on orthostatic performance in school and other daily activities and can be treated by various non-pharmaceutical and pharmaceutical interventions.

### Reported Lack of Medical Care

Although ME/CFS is defined as a disorder that significantly reduces or impairs daily activities, only one of our patients had previously received a certificate of disability and none was supported by adequate medical devices or home care, together reflecting poor medical care and barriers to specialized support as outlined in previous studies [67–69]. A large proportion of our patients took various dietary supplements and/or had received complementary medical treatment which both had also been demonstrated for other pediatric and adult ME/CFS cohorts and may, in addition, reflect a lack of standard medical support [7, 92]. The lack of adequate medical care is especially remarkable regarding the large number of medical consultations with ME/CFS symptoms prior to diagnosis reported by our patients and in other studies on ME/CFS [67, 69].

All pupils in our study reported on frequent school absences, and, remarkably, only a minority had received any educational assistance such as home or digital schooling. These findings were in line with earlier studies indicating that ME/CFS is a frequent cause of prolonged school absences and severely affects social participation and education of young people [3–8]. This is particularly concerning, since pediatric patients with ME/CFS reported that remaining engaged in an education system that flexibly accommodated their illness and aspirations was crucial for their long-term functioning [7, 93, 94].

### Laboratory Findings

So far, no biomarker for ME/CFS has been established. Symptom-oriented laboratory analyses are recommended to evaluate potential differential diagnoses and usually show unremarkable results in ME/CFS [6, 54]. This was also true for our cohort. Some patients demonstrated minor deviations of hematological and immunological parameters, including mild anemia, lymphocytosis, and/or neutropenia as well as mild changes in peripheral lymphocyte subtypes and serological markers of inflammation, autoimmunity, and/or allergies. More than half of the patients showed elevated ANA titers, as previously reported for many ME/CFS patients, without clear evidence of distinct underlying autoimmune diseases [7]. The proportion of positive titers (56%) was greater than expected from a previous report with 34% positive titers in this age group [7]. Although, it is not unknown to have moderately elevated titers without evidence of autoimmune disorders in pediatric rheumatology. Almost a third had elevated IgE levels but clear associations between IgE levels and ME/CFS have not been found in earlier studies [95]. Remarkably, the majority of patients presented with vitamin D deficiency. However, low serum concentrations of vitamin D did not appear to be a contributing factor to the level of fatigue in another cohort of ME/CFS patients [96].

As expected, all patients showed anti-EBV VCA IgG as an indicator of previous EBV infection, with some individuals lacking an anti-EBNA-1 seroconversion. The findings of undetectable EBNA- 1 IgG as well as of detectable anti-EBV VCA IgM, anti-EBV EA IgG, EBV DNA in throat washes were not more frequent in our cohort than reported for the general population [97–100]. Detectable EBV DNA in blood cells was more frequent than in a U.S. cohort aged 1 – 97 years (median 42 years) without EBV-associated disorder [101]. In sum, we found no significant correlation between EBV-specific serological findings or detection of EBV DNA with disease severity or physical functioning. These findings were in line with previous research that has not found a distinct pattern of EBV-specific routine virological results in ME/CFS patients [42, 102]. However, findings regarding EBV-specific immune responses in patients with ME/CFS are inconsistent, and EBV antigen mimicry might well contribute to pathogenic autoimmunity as shown by us in more comprehensive EBV-specific immunological analyses [35, 44, 46, 103–107]. EBV and herpes viruses in general have been discussed as a cause or perpetuating factor of ME/CFS, while no stringent causal association has been proven yet [41, 42, 102].

### Partial Remission at Follow-up

In a large Australian cohort aged 16 to 77 years, post-infectious fatigue triggered by EBV, Q-fever or Ross River virus was transient in most cases, as indicated by 35%, 27%, 12%, and 9% of patients affected at 1.5, 3, 6, and 12 months after the onset of symptoms [19]. However, most concerningly, an increasing number of studies on long-term sequelae of coronavirus disease 2019 (COVID 19) currently indicate that most patients affected by post-COVID syndrome at 3 months were still not recovered after 12 months [22, 108].

Most studies specifically assessing fatigue and ME/CFS in young people after EBV-IM evaluated no longer follow-up periods than 6 months, some relied on self-reported ME/CFS or did not require PEM for ME/CFS diagnosis, and none compared the course of disease in adolescents and very young adults [19, 20, 24–31]. However, according to a more favorable prognosis in younger people, the proportion of ME/CFS after EBV-IM in adolescents in the US decreased significantly over time, with 12.7%, 7%, and 4% at 6, 12, and 24 months [18, 19].

In our study the median time since onset of EBV-IM was 13.8 months at the first visit, indicating that some patients might recover over time during the study period. Most remarkably, the majority of our adolescent patients did not fulfill the criteria for ME/CFS diagnosis any more at 12 months after the first visit, while all adult patients still met the CCC. The different health trajectories for adolescent and adult patients were also evident in the self-perceived health transition item of the SF- 36 at 12 months after the first visit, with 40% and 20% of children rating their general health as much better or somewhat better, and 45% and 22% of adults much worse or somewhat worse than in the previous year, respectively. Furthermore, over the whole study period symptom load (see below) and school functioning (PedsQL) significantly improved in adolescents while remaining stable or worsening in adults.

These findings were in line with compelling evidence indicating a better ME/CFS prognosis of children and adolescence compared to adults, with pediatric studies reporting recovery of up to 83% [6–8, 15–17, 27, 54–61]. A large pediatric follow-up analysis from Australia indicated recovery in 50% of the cases at five and 68% of the cases at ten years after disease onset with a median disease duration of five (range 1-15) years [7]. Dramatic improvement was reported to be more likely within the first four years [6]. Accordingly, all patients in partial remission in our cohort had an illness duration of less than three years. Although some adult patients, in general, may also experience significant improvements over time, a systematic review indicated that prognosis in adults is fairly poor, with only a minority of adult patients experiencing full recovery [109].

Importantly, only two of our pediatric patients were largely free of ME/CFS symptoms (except OI) at their last visit and all remaining patients, independently of their state of partial remission, still reported symptoms from several domains of the diagnostic criteria. Furthermore, we observed some fluctuation of disease load over time, with some patients no longer meeting the diagnostic criteria at 6 but again at 12 months, thereby reflecting unstable and unpredictable dynamics of remissions. Remissions and relapses were reported to be common in pediatric ME/CFS patients and can occur for example after overexertion or additional infectious illnesses [6]. Our findings support the recommendation that pediatric patients must be offered thorough follow-up evaluations and continuous medical care even if they do not any longer fulfill the diagnostic ME/CFS criteria.

It remains challenging to measure recovery from ME/CFS, especially in pediatric patients. Previous pediatric studies have established recovery through various measures such as school attendance, fulfillment of diagnostic criteria, or self-report [55]. However, available studies have used various diagnostic criteria, different follow-up periods (ranging from six months to 13 years), and different inclusion criteria, thereby limiting direct comparisons. Moreover, what young people with ME/CFS consider as “recovery” can largely differ [7] and patient coping at any age might challenge measuring recovery [110].

Risk factors for prolonged disease after EBV-IM and for ME/CFS duration in general are not yet clear. US college students who developed ME/CFS after IM had more physical complaints but not more perceived stress, depression, anxiety, or abnormal coping before the onset of IM [20]. Some studies have associated prolonged symptoms after EBV-IM with older age and severity of acute IM, others have identified initial pain severity, autonomic symptoms, lower mental health scores, higher anxiety and depression scales, more days-in-bed, female gender, and distinct laboratory changes (CRP, vitamin B12, cytokines) during EBV-IM as risk factors for the development of ME/CFS. However, the case definitions of ME/CFS varied in early studies, findings have been inconsistent, and it is unknown, whether risk factors of ME/CFS onset also impact on the duration of ME/CFS [20, 24, 28–30, 47, 111]. Several factors have been suggested to affect the prognosis of ME/CFS in general, including age, female gender, fatigue severity at disease onset, PEM severity, severity of ME/CFS symptoms, comorbidities, illness duration, life stressors, and lower socioeconomic status [6, 15, 16, 84, 112, 113], although findings so far remained inconclusive. With better initial test results and better final outcomes of adolescents compared to adults, our findings might support younger age, shorter disease duration, a better Bell Score, as well as less severe fatigue (CFQ) at first presentation as potential markers for a more favorable course of disease although, of course, the number of patients was by far too small to draw any final conclusions.

The interpretation of published data on ME/CFS outcome is additionally challenged by the fact that in many ME/CFS cohorts the initial trigger is less well characterized than in our cohort. An Australian ME/CFS cohort included 80% post-infectious cases, with 40% of the cases described as triggered by EBV-IM, although only 25% had documented serological changes at onset [7]. Other pediatric studies have reported between 23% and 90% of perceived infectious triggers [15–17, 114] or provide no further information on any events at onset [8, 27, 56]. Some studies, furthermore, included patients with dominantly gradual onset and thus no distinct trigger [5, 17].

### Symptom Load over the Study Period

Our patients reported a wide range of symptoms that persisted over time, with overall little change in severity or frequency. The ME/CFS core symptoms PEM, fatigue, need for rest, performance limitations in daily life, unrefreshing sleep, concentration problems, and mental exhaustibility were reported to occur at least frequently and to be at least severe by a substantial proportion of patients. The results of this study highlight the interindividual variability of dominant symptoms as well as the fluctuation of intraindividual symptoms over the course of a year. In general, symptoms in pediatric ME/CFS are reported to fluctuate more than in adults [6]. However, due to the small size of our cohort we did not investigate this issue in more detail.

In adolescent patients, fewer symptoms were reported at 6 and 12 months compared to the initial visit, which is in line with partial remission of some patients. Conversely, the number of symptoms reported by adults remained stable over the whole study period. It is also worth noting that adults consistently reported more symptoms and almost twice as many frequent symptoms than adolescents at all visits. This supports the recommendation to quantify symptoms at the time of diagnosis and might suggest prioritizing frequent symptoms in treatment protocols. Quantifying frequency and specificity was recommended to increase the specificity of ME/CFS diagnosis [115], since mild forms of most ME/CFS symptoms frequently occur in the general population. In our cohort a substantial number of symptoms were also reported to occur only sometimes or rarely, or to be of mild severity.

### Health-Related Quality of Life

Previous studies have shown that children and young adults with ME/CFS experience a severe illness that fundamentally affects their social life and education, and is associated with a poorer HRQoL than found in peers with other chronic diseases including asthma, diabetes mellitus, epilepsy, and cystic fibrosis [7, 9–11]. Remarkably, the PedsQL results were very similar between our adolescent cohort and adolescent ME/CFS patients in the US, Australia or Norway [9, 10, 116, 117]. Furthermore, our adolescent sample also displayed the same relative distribution of subscales as published cohorts, with patients reporting worse HRQoL in physical and school function and better HRQoL in social and emotional functioning.

Adolescents in our cohort reported moderate improvements on the total score, physical and psychosocial score, as well as significant improvements on the school domain, while social and emotional quality of life seemed stable. The improvements in the total score, psychosocial score, and school score were greater than a suggested minimal clinically meaningful difference in PedsQL results from pediatric cohorts [118]. However, no clinically meaningful difference was specifically suggested so far for patients with ME/CFS. The improvements observed in HRQoL of our adolescent cohort are in line with the fact that some of the patients experienced a partial remission. However, intensive school counselling by our team might have contributed to a better school situation and changes in HRQoL.

Most concerningly, we found little evidence for improved HRQoL in young adults, apart from improvements in the emotional and social subdomains possibly due to the implementation of specialized psychosocial and medical care. Young adults in our cohort reported much lower HRQoL compared to adolescents from our and other studies. We are not aware of any other studies assessing HRQoL in very young adults specifically, but studies in adult ME/CFS patients in general have consistently reported very low HRQoL [119, 120].

The transition from adolescence to young adulthood as well as from pediatrics to adult patient medicine can be particularly challenging for young people with ME/CFS, with individuals facing a range of uncertainties regarding health care, education, financials, and contact to peers. These factors may be contributing to the lower HRQoL reported by young adults compared with adolescents in our study. Unrevealing the additional age-specific risk factors for chronification will be crucial for developing effective strategies to improve the well-being of this most vulnerable group of people with post-infectious syndromes and ME/CFS.

Few studies have investigated risk factors for worse HRQoL in adolescents with ME/CFS and found high frequency of PEM, cognitive symptoms, regular school absence, delayed school progression, and attending physical therapy or rehabilitation as possible risk factors. After diagnosis, support of school personal, regular school attendance, and participation in leisure activities have been associated with higher HRQoL [10, 121]. The findings regarding a possible risk contribution of depressive symptoms were inconsistent [10, 117, 120, 121]. Furthermore, it was reported that adolescents meeting the Fukuda and IOM criteria displayed worse HRQoL than adolescents only satisfying the Fukuda criteria, suggesting that criteria requiring PEM might select patients with worse HRQoL [10]. The fact that the very strict CCC have been used to diagnose ME/CFS in our adult patients might have contributed to the findings of lower HRQoL, worse physical functioning, and longer disease duration in this age group. Additionally, a selection bias must be considered, as the sample of young adults in this study may have been skewed by the fact that they sought treatment at a pediatric clinic, potentially indicating a higher level of desperation or urgency in seeking medical care.

Altogether, it is difficult to interpret the trajectories of HRQoL in this study, as to our best knowledge, no other longitudinal evaluations of HRQoL in children or young adults with ME/CFS are available, and since no matched healthy controls have been evaluated during the study period. Especially a possible impact of fears elicited by the COVID-19 pandemic or by restrictions thereof during the course of disease cannot be excluded.

### Strengths and Limitations

As an important strength our study presents longitudinal data on ME/CFS cases that manifested after serologically confirmed EBV-IM. Confirming an infectious onset or even a specific infectious trigger of ME/CFS years after its occurrence can be very challenging, since self-reports are not reliable and ordering former medical documents is time-consuming, and in many cases not successful. This is especially true in cases of EBV-IM, where retrospective diagnoses rely on sufficient serological evidence of EBV primary infection documented together with typical clinical signs and symptoms. We did not include patients with probable EBV-IM (e.g. typical clinical signs only) in this article which we will report on together with a young cohort of ME/CFS patients triggered by other infectious diseases. Our study supports the reassurance of younger adolescents that improvement of ME/CFS following EBV-IM is not uncommon, even if it takes months or years, and that remission is possible, also it cannot be guaranteed, as suggested by others [6, 7, 18].

A second strength of our study is the inclusion of very young adult patients together with adolescents. The former population often gets lost from pediatric as well as non-pediatric studies due to age-specific challenges on either side, including studies on post-viral syndromes. For example, a recent longitudinal study on long-term recovery from SARS-CoV-2 infection in children specifically excluded participants from follow-up after their 18^th^ birthday and thereby possibly generated a bias towards lower prevalence of complex and severe symptoms [122]. For patients older than 17 years pediatricians mostly cannot provide follow-up due to regulatory issues, and non-pediatric care takers might face barriers to getting a complete medical history and/or to helping with issues regarding school, academic career, parents, siblings, or peers. Unfortunately, we witnessed difficulties of very young adults with ME/CFS to be adopted by local general practitioners which most likely further impaired their clinical outcome. In our center we regularly see young people up to an age of 20 to facilitate the transition within age-specific borders of medical care and to investigate age-specific resilience and risk factors.

Third and importantly, we provide data on ME/CFS cases that were diagnosed by clinical criteria requiring PEM as a hallmark of ME/CFS and as recommended by the EUROMENE [48] and others [49, 50, 52]. Moreover, we compare the fulfillment of the CCC and the less strict criteria of the CDW-R in our adolescent cohort, while many earlier studies reported on EBV-triggered ME/CFS cases that were diagnosed by the polythetic Fukuda or even the Oxford criteria which both do not essentially require PEM.

We furthermore provide detailed information on clinical phenotypes, including the frequency and severity of typical ME/CFS symptoms, a comprehensive set of data from various well-established PROMs evaluating fatigue, daily functioning, or HRQoL (CFQ, Bell Score, PedsQL, SF-36), and various laboratory results, including peripheral immunophenotypes, EBV serology, and EBV DNA load.

Overall, our study adds to the current understanding of ME/CFS in young people and highlights the importance of an early diagnosis as well es of a thorough longitudinal evaluation of patients with ME/CFS following EBV-IM.

The study has several limitations that must be considered when interpreting the results. First, a potentially multi-facetted selection bias is limiting the generalizability of results. Since our center is the only specialized pediatric ME/CFS clinic in Germany, before admission patients are faced with significant waiting times and with time-consuming prerequisites such as answering questionnaires and providing previous medical results. Additionally, many patients must travel a significant distance to reach our clinic and therefore the study sample is most likely skewed towards individuals with moderate or less severe ME/CFS and to families than can afford travelling. Moreover, our cohort was limited to children and adolescents registered in upper school services and to Caucasian patients, which might suggest that individuals from other groups may be underrepresented, possibly due to barriers within the health care system; given the lack of knowledge on ME/CFS, it often requires significant effort of patients and their families to get specialized care and not rarely the patients’ journeys involve multiple healthcare professionals.

Second, the sample size of the study and the number of individuals who experienced partial remission is relatively small, which can affect the statistical power of the analyses and the generalizability of the results.

Third, although the drop-out rate of 20% at the assessment after 12 months was deemed acceptable, it is important to acknowledge that some patients were lost to follow-up, which could introduce bias and potentially reduce the overall representativeness of our sample. Based on our experience, discontinuation in ME/CFS studies may not be missing at random, as some patients may be unwilling to continue participating due to perceived improvement, while others may feel too ill to continue.

In addition, this study was limited in its ability to investigate risk factors prior to diagnosis of ME/CFS, since patients were registered after the onset of this disease. Our study focused on examining risk factors that might have contributed to the maintenance of ME/CFS. However, it is important to note that additional prospective studies, such as those in US college students, are needed to further explore potentially predisposing factors indicating an increased risk or resilience without recall bias [20].

As another limitation it should be noted that the 12 months follow-up period may not provide enough information to evaluate the long-term prognosis of ME/CFS, particularly in young adults. Longer follow-up data would be beneficial, especially since young adults do not appear to show significant improvement one year after diagnosis.

Finally, the lack of a matched control group in this study makes it challenging to interpret some results, for example changes in HRQoL over time or frequency and severity of ME/CFS symptoms that are non-specific and common in the general population.

While this study provides valuable insights into the clinical course of a complex chronic disease in young people, these limitations suggest that caution is necessary when interpreting the results. Future studies with larger sample sizes, longer follow-up periods, and appropriate control groups are necessary to further validate and extend these findings.

## Conclusions

In conclusion, ME/CFS after EBV-IM is a debilitating disease that results in severe functional impairment and poor quality of life for both adolescents and young adults in Germany. Access to healthcare and treatment is a fundamental barrier for patients with this condition. Laboratory findings were inconclusive, and there was no evidence to suggest that EBV perpetuates the disease. While patients with ME/CFS often experience a variety of fluctuating symptoms, it is possible that the frequency and severity of core symptoms may be more significant in predicting prognosis. Young adults reported more symptoms, greater physical impairment, and worse quality of life than adolescents. While some pediatric patients showed signs of improvement over the study period, young adults tended to have a poor prognosis, underscoring the need for special attention when taking care for this age group. There is a clear need for more research to clarify the reasons for the different health trajectories and to identify prognostic markers that allow for an evidence-based stratification of multi-professional and multi-modal longitudinal care. Further research is also necessary to better understand the pathomechanisms of the disease, identify reliable biomarkers for early diagnosis, and to develop effective treatments that can improve outcomes and support recovery.

## Data Availability Statement

Data is available upon reasonable request.

## Ethics Statement

Patients and parents (of patients < 18 years) gave written and informed consent prior to inclusion. The study was approved by the local Ethics Committee (529/18, 485/18) and conducted in accordance with the Declaration of Helsinki.

## Author Contributions

R.P., P.M., T.H., K.G., and U.B. designed the study. R.P., P.M., T.H., H.Z., Y.M., K.W, J.P., and A.L. acquired data and investigated patients. R.P., S.M.H., L.M, and U.B. analyzed the data. R.P., M.H., L.M., and U.B. drafted and reviewed the manuscript. K.G. and U.B. supervised the work. All authors discussed the results and commented on the manuscript.

## Funding

This work has been supported by the Lost Voices and Weidenhammer-Zoebele foundations.

## Acknowledgments

We thank all patients who participated in this study and their parents for supporting their participation.

## Conflict of Interest

U.B. received research grants from Federal Ministry of Education and Research (BMBF), the Federal Ministry of Health (BMG), the Bavarian Ministry of Health and Care (StMGP), the Bavarian Ministry of Science and Arts (StMWK), the German Center for Infection Research (DZIF), the People for Children (Menschen für Kinder) Foundation, the Weidenhammer-Zöbele Foundation, the Lost-Voices Foundation, and the ME/CFS Research Foundation.

C.S. was consulting Roche, Celltrend, and Bayer; she received support for clinical trials by Bayer, Fresenius, and Miltenyi, honoraria for lectures by Fresenius, AstraZeneca, BMS, Roche, Bayer, and Novartis, and research grants from the German Research Association (DFG), the BMBF, the BMG, the Weidenhammer-Zoebele Foundation, the Lost-Voices Foundation, and the ME/CFS Research Foundation.

All other authors declare that the research was conducted in the absence of any commercial or financial relationships that could be construed as a potential conflict of interest.

## Notes

### Funding Statement

This study was funded by the Lost Voices and Weidenhammer-Zoebele foundations.

### Author Declarations

Ethics committee/IRB of the Technical University of Munich gave ethical approval for this work (529/18, 485/18).

