## Supplementary Material for "One-Year Follow-up of Young People with ME/CFS Following Infectious Mononucleosis by Epstein-Barr Virus"

### 1 Supplementary Figures and Tables

#### 1.1 Supplementary Figures

**Supplementary Figure S1.** Time-to-event analysis of adolescent and adult patients from onset of Epstein-Barr virus infectious mononucleosis (EBV-IM) to diagnosis of myalgic encephalomyelitis/chronic fatigue syndrome (ME/CFS) following presentation to the MRI Chronic Fatigue Center (MCFC). The x-axis represents the time (in days) from onset of EBV-IM, while the y-axis represents the proportion of patients who received a diagnosis of ME/CFS at the MCFC. The shaded areas represent the 95% confidence band. There was no significant difference in time-to-diagnosis at the MCFC between adolescents and adult patients (P > 0.05).

#### 1.2 Supplementary Tables

#### Supplementary Table S1. Immunophenotyping of Peripheral Blood Lymphocytes

| **Laboratory result** | **All^1^** | **Adult^1^** | **Adolescents^1^** | **p-value^2^** |
| --- | --- | --- | --- | --- |
| **Pan-T cells** (absolute) | | | | 0.604 |
| Within normal range | 19/23 (83) | 10/13 (77) | 9/10 (90) |  |
| Above normal range | 4/23 (17) | 3/13 (23) | 1/10 (10) |  |
| Below normal range | 0/23 (0) | 0/13 (0) | 0/10 (0) |  |
| **Pan-T cells** (relative) | | | | 0.178 |
| Within normal range | 21/23 (91) | 13/13 (100) | 8/10 (80) |  |
| Above normal range | 2/23 (9) | 0/13 (0) | 2/10 (20) |  |
| Below normal range | 0/23 (0) | 0/13 (0) | 0/10 (0) |  |
| **Pan-B cells** (absolute) | | | | 0.771 |
| Within normal range | 18/23 (78) | 11/13 (85) | 7/10 (70) |  |
| Above normal range | 2/23 (9) | 1/13 (8) | 1/10 (10) |  |
| Below normal range | 3/23 (13) | 1/13 (8) | 2/10 (20) |  |
| **Pan-B cells** (relative) | | | | 0.178 |
| Within normal range | 21/23 (91) | 13/13 (100) | 8/10 (80) |  |
| Above normal range | 0/23 (0) | 0/13 (0) | 0/10 (0) |  |
| Below normal range | 2/23 (9) | 0/13 (0) | 2/10 (20) |  |
| **NK cells** (absolute) | | | |  |
| Within normal range | 22/22 (100) | 13/13 (100) | 9/9 (100) |  |
| Above normal range | 0/22 (0) | 0/13 (0) | 0/9 (0) |  |
| Below normal range | 0/22 (0) | 0/13 (0) | 0/9 (0) |  |
| **NK cells** (relative) | | | | >0.999 |
| Within normal range | 21/22 (95) | 12/13 (92) | 9/9 (100) |  |
| Above normal range | 0/22 (0) | 0/13 (0) | 0/9 (0) |  |
| Below normal range | 1 (5) | 1/13 (8) | 0/9 (0) |  |
| **HLA-DR+ cells** (absolute) | | | | 0.245 |
| Within normal range | 15/18 (83) | 11/12 (92) | 4/6 (67) |  |
| Above normal range | 3/18 (17) | 1/12 (9) | 2/6 (33) |  |
| Below normal range | 0/18 (0) | 0/12 (0) | 0/6 (0) |  |
| **HLA-DR+ cells** (relative) | | | | 0.147 |
| Within normal range | 18/20 (90) | 12/12 (100) | 6/8 (75) |  |
| Above normal range | 1/20 (5) | 0/12 (0) | 1/8 (12) |  |
| Below normal range | 1/20 (5) | 0/12 (0) | 1/8 (12) |  |

^1^ n (with positive result) / n (investigated) (%)

^2^ Fisher’s exact test

NK, Natural killer cells; HLA-DR, Human Leukocyte Antigen DR

**Supplementary Table S2.** Comparison of Recovered and Non-recovered Patients

| Characteristics | Non-recovered | Recovered | p-value^1^ |
| --- | --- | --- | --- |
| Number of patients | n=19 | n=6 |  |
| Age group and illness duration | | | |
| Adolescents^4^ | 6/19 (32) | 6/6 (100) | **0.005** |
| Illness duration in months from onset to first visit^2^ | 16.9 (9.6 – 41.8) | 10.8 (6.7 – 15.4) | 0.503 |
| Baseline questionnaire results | |  |  |
| Chalder fatigue scale^3^ | 26 (5) | 21 (1) | **0.001** |
| Bell Score^2^ | 30 (30 – 40) | 50 (43 – 50) | 0.054 |
| SF-36 PCS^3^ | 28 (8) | 31 (10) | 0.562 |
| SF-36 MCS^3^ | 40 (11) | 49 (7) | 0.074 |
| PedsQL^3^ | 42 (13) | 56 (11) | **0.026** |
| Gender^4^ |  |  |  |
| Female | 16/19 (84) | 4/6 (67) | 0.562 |
| ME/CFS criteria^4^ |  |  |  |
| CCC | 16/19 (84) | 5/6 (83) | >0.999 |
| CDW-R | 6/7 (86) | 6/6 (100) | >0.999 |
| Comorbidity^4^ |  |  |  |
| PoTS | 16/19 (84) | 3/4 (75) | >0.999 |
| Allergies | 10/19 (53) | 2/6 (33) | 0.645 |
| Current medical care^4^ |  |  |  |
| Nutrition supplements | 14/18 (78) | 3/6 (50) | 0.131 |
| Prescription medication | 9/18 (50) | 2/6 (33) | 0.348 |
| Family history^4^ |  |  |  |
| ME/ CFS in family | 2/19 (11) | 0/6 (0) | >0.999 |
| AID in family | 8/19 (42) | 2/6 (33) | >0.999 |
| IM in family | 12/19 (63) | 4/6 (69) | >0.999 |
| Selected laboratory parameter^4^ | | | |
| ANA ↑ | 11/19 (58) | 3/6 (50) | >0.999 |
| IgE ↑ | 5/19 (26) | 2/6 (33) | >0.999 |
| 25-OH-Vitamin-D ↓ | 10/19 (56) | 4/6 (67) | >0.999 |
| ^1^ Fisher's exact test; Wilcoxon rank sum test; Pearson's Chi-squared test | | | |
| ^2^ Median (IQR) ^3^ Mean (SD)  ^4^ Number of patients with indicated characteristic/number of patients investigated (%)  ↑ above normal range; ↓ below normal range;  CCC, Canadian Consensus Criteria [49], CDW-R, Clinical Diagnostic Worksheet [6]; PoTS, postural orthostatic tachycardia syndrome; AID, autoimmune disease; IM, infectious mononucleosis; PedsQL, Pediatric Quality of Life Inventory; SF-36 PCS, Short Form 36 Health Survey Physical Component Summary Score; SF-36 MCS, Short Form 36 Health Survey Mental Health Component Summary Score. ANA, antinuclear antibodies; | | | |

**Supplementary Table S3**: EBV Serology, PCR and IgG Immunoblot of Recovered and Non-recovered Patients

| **EBV Diagnostics** | **Non-recovered**   n/n (%)^1^ | **Recovered**  n/n (%)^1^ | **p-value**^2^ |
| --- | --- | --- | --- |
| EBV PCR |  |  |  |
| DNA in cell fraction |  |  | 0.535 |
| – | 10/16 (63) | 2/4 (50) |  |
| (+) | 4/16 (25) | 1/4 (25) |  |
| + | 2/16 (12) | 1/4 (25) |  |
| DNA in plasma |  |  |  |
| – | 19/19 (100) | 6/6 (100) |  |
| + | 0/19 (0) | 0/6 (0) |  |
| DNA in throat wash |  |  | 0.452 |
| – | 9/19 (47) | 2/6 (33) |  |
| + | 10/19 (53) | 4/6 (67) |  |
| EBV ELISA |  |  |  |
| VCA IgM |  |  | 0.451 |
| – | 12/19 (63) | 0/6 (0) |  |
| (+) | 1/19 (6) | 0/6 (0) |  |
| + | 6/19 (32) | 6/6 (100) |  |
| VCA IgG |  |  |  |
| – | 0/19 (0) | 0/6 (0) |  |
| + | 19/19 (100) | 6/6 (100) |  |
| EBNA1 IgG |  |  | 0.430 |
| – | 1/19 (5) | 1/6 (17) |  |
| + | 18/19 (95) | 5/6 (83) |  |
| EBV IgG Immunoblot |  |  |  |
| EAp54 |  |  | 0.533 |
| – | 11/19 (58) | 3/6 (50) |  |
| (+) | 5/19 (26) | 1/6 (17) |  |
| + | 3/19 (16) | 2/6 (33) |  |
| EAp138 |  |  | 0.676 |
| – | 11/19 (58) | 3/6 (50) |  |
| (+) | 5/19 (26) | 1/6 (17) |  |
| + | 3/19 (16) | 2/6 (33) |  |
| BZLF1 |  |  | 0.793 |
| – | 3/19 (16) | 0/0 (0) |  |
| (+) | 3/19 (16) | 1/6 (17) |  |
| + | 13/19 (68) | 5/6 (83) |  |
| VCAp23 |  |  | >0.999 |
| – | 2/19 (11) | 0/6 (0) |  |
| (+) | 0/19 (0) | 0/6 (0) |  |
| + | 17/19 (89) | 6/6 (100) |  |
| VCAp18 |  |  | >0.999 |
| – | 1/19 5) | 0/6 (0) |  |
| (+) | 0/19 (0) | 0/6 (0) |  |
| + | 18/19(95) | 6/6 (100) |  |
| EBNA-1 |  |  | 0.579 |
| – | 1/19 (5) | 1/6 (17) |  |
| (+) | 1/19 (5) | 0/6 (0) |  |
| + | 17/19 (89) | 5/6 (83) |  |
| ^1^ Number of patients with indicated laboratory parameter/ number of patients investigated (%)  ^2^ Fisher’s exact test | | | |
| EBV, Epstein-Barr Virus; VCA, virus capsid antigen; EBNA, EBV nuclear antigen; EA, early antigen. | | | |

**Supplementary Table S4.** Symptom Burden over the Study Period

|  | All (n=25) | | | | Adults (n=13) | | | | Adolescents (n=12) | | | |
| --- | --- | --- | --- | --- | --- | --- | --- | --- | --- | --- | --- | --- |
| Number of symptoms | First Visit^1^ | 6 Months^1^ | 12 Months^1^ | p-value^2^ | First Visit^1^ | 6 Months^1^ | 12 Months^1^ | p-value^2^ | First Visit^1^ | 6 Months^1^ | 12 Months^1^ | p-value^2^ |
| All | 27 (5) | 25 (7) | 25 (9) | 0.487 | 29 (4) | 27 (6) | 29 (4) | 0.896 | 25 (7) | 22 (8) | 21 (10) | 0.759 |
| At least sometimes | 23 (6) | 21 (8) | 21 (9) | 0.895 | 26 (4) | 25 (7) | 27 (5) | 0.772 | 19 (4) | 17 (8) | 16 (9) | 0.819 |
| At least frequently | 15 (5) | 14 (8) | 14 (8) | 0.723 | 19 (5) | 19 (6) | 20 (5) | 0.785 | 12 (3) | 10 (6) | 9 (6) | 0.306 |
| Always | 5 (4) | 4 (4) | 5 (5) | 0.836 | 7 (4) | 6 (4) | 8 (5) | 0.716 | 2 (1) | 3 (3) | 2 (2) | 0.952 |
| ^1^ Mean (SD)  ^2^ Kruskal-Wallis rank sum test | | | | | | | | | | | | |

**Supplementary Table S5.** Difference in Symptom Burden Between Adolescents and Young Adults

|  | **First Visit** | | | **6 Months** | | | **12 Months** | | |
| --- | --- | --- | --- | --- | --- | --- | --- | --- | --- |
| Number of symptoms | Adults^1^ | Adolescents^1^ | p-value^2^ | Adults^1^ | Adolescents^1^ | p-value^2^ | Adults^1^ | Adolescents^1^ | p-value^2^ |
| All | 29 (3) | 25 (7) | 0.084 | 27 (6) | 22 (8) | 0.051 | 29 (4) | 21 (10) | **0.041** |
| At least sometimes | 26 (4) | 19 (4) | **<0.001** | 25 (7) | 17 (8) | **0.030** | 27 (4) | 16 (9) | **0.004** |
| At least frequently | 19 (5) | 12 (3) | **0.006** | 19 (6) | 10 (6) | **0.023** | 20 (5) | 9 (7) | **0.002** |
| Always | 7 (4) | 2 (1) | **0.009** | 5 (4) | 3 (3) | 0.059 | 8 (5) | 3 (3) | **0.012** |
| ^1^ Mean (SD)  ^2^ Wilcoxon rank sum test | | | | | | | | | |

**Supplementary Table S6.** Differences in Patient-reported Outcome Measures Between Adolescents and Young Adults

|  | **First Visit** | | | **6 Months** | | | **12 Months** | | |
| --- | --- | --- | --- | --- | --- | --- | --- | --- | --- |
|  | Adults | Adolescents | p-value^1^ | Adults | Adolescents | p-value^1^ | Adults | Adolescents | p-value^1^ |
| Chalder fatigue^2^ scale | 28 (4) | 22 (5) | **0.006** | 28 (4) | 19 (9) | **0.016** | 29 (4) | 18 (9) | **0.003** |
| Bell Score^3^ | 30 (30 – 40) | 50 (50 – 50) | **0.019** | 30 (30 – 40) | 60 (40 – 80) | **0.019** | 30 (25 – 40) | 60 (40 – 80) | **0.007** |
| PedsQL^2^ | | | | | | | | | |
| Total Score | 35 (11) | 54 (9) | **0.002** | 37 (15) | 62 (14) | **0.003** | 37 (15) | 61 (20) | **0.018** |
| Physical | 30 (12) | 48 (18) | **0.035** | 29 (13) | 60 (22) | **0.003** | 25 (15) | 53 (27) | **0.017** |
| Social | 50 (18) | 77 (16) | **0.007** | 49 (15) | 76 (16) | **0.003** | 51 (15) | 76 (16) | **0.007** |
| Emotional | 34 (15) | 61 (17) | **0.004** | 36 (23) | 60 (19) | **0.015** | 44 (21) | 60 (27) | 0.127 |
| School | 29 (18) | 35 (11) | 0.517 | 37 (22) | 48 (23) | 0.246 | 33 (26) | 58 (25) | 0.087 |
| Psychosocial | 38 (13) | 58 (6) | **0.001** | 41 (17) | 63 (14) | **0.011** | 45 (19) | 64 (18) | **0.030** |
| SF-36^2^ | | | | | | | | | |
| Physical functioning | 42 (21) | 65 (25) | **0.039** | 46 (20) | 71 (25) | **0.039** | 35 (27) | 73 (31) | **0.019** |
| Role physical | 7 (12) | 17 (16) | 0.129 | 3 (8) | 30 (40) | 0.083 | 6 (11) | 38 (46) | 0.129 |
| Pain | 34 (21) | 47 (29) | 0.235 | 34 (15) | 62 (29) | **0.039** | 37 (23) | 60 (31) | 0.091 |
| General health | 22 (11) | 30 (12) | 0.133 | 21 (9) | 35 (15) | **0.032** | 20 (7) | 37 (20) | 0.052 |
| Vitality | 15 (13) | 29 (12) | **0.012** | 18 (11) | 38 (24) | 0.050 | 11 (8) | 42 (28) | **0.010** |
| Social | 31 (23) | 51 (27) | 0.085 | 24 (22) | 59 (32) | **0.025** | 24 (25) | 58 (36) | 0.051 |
| Role Emotional | 64 (46) | 78 (33) | 0.561 | 52 (44) | 58 (40) | 0.844 | 56 (47) | 73 (41) | 0.475 |
| Mental health | 52 (20) | 65 (17) | 0.121 | 43 (20) | 63 (19) | **0.025** | 51 (18) | 66 (21) | 0.093 |
| Physical component summary score | 26 (8) | 32 (9) | 0.151 | 28 (8) | 38 (13) | 0.056 | 25 (9) | 38 (13) | **0.013** |
| Mental health component summary score | 39 (12) | 45 (9) | 0.211 | 34 (11) | 42 (14) | 0.261 | 37 (11) | 45 (12) | 0.182 |
| ^1^Wilcoxon rank sum test.  ^2^ Mean (SD)  ^3^ Median (IQR)  PedsQL, Pediatric Quality of Life Inventory; SF-36, Short Form 36 Health Survey. | | | | | | | | | |

**Supplementary Table S7.** Self-perceived Change of General Health Status over Time

| General health status  compared to previous year | First Visit | 12 Months |
| --- | --- | --- |
| **All** | n=23 | n=19 |
| Much worse | 8 (35%) | 4 (21%) |
| Somewhat worse | 5 (22%) | 3 (14%) |
| About the same | 5 (22%) | 6 (32%) |
| Somewhat better | 4 (17%) | 2 (10%) |
| Much better | 1 (4%) | 4 (21%) |
| **Adolescents** | n=12 | n=10 |
| Much worse | 3 (25%) | 0 (0%) |
| Somewhat worse | 5 (42%) | 1 (10%) |
| About the same | 3 (25%) | 3 (30%) |
| Somewhat better | 1 (8%) | 2 (20%) |
| Much better | 0 (0%) | 4 (40%) |
| **Adults** | n=11 | n=9 |
| Much worse | 5 (45%) | 4 (44%) |
| Somewhat worse | 0 (0%) | 2 (22%) |
| About the same | 2 (18%) | 3 (33%) |
| Somewhat better | 3 (27%) | 0 (0%) |
| Much better | 1 (9%) | 0 (0%) |
